# Continuity of routine immunization programs in Canada during the COVID-19 pandemic

**DOI:** 10.1101/2021.03.22.21254121

**Authors:** Hannah Sell, Ali Assi, S. Michelle Driedger, Eve Dubé, Arnaud Gagneur, Samantha B. Meyer, Joan Robinson, Manish Sadarangani, Matthew Tunis, Shannon E. MacDonald

**Author notes:** **Corresponding author**: Shannon MacDonald, Mailing address: Faculty of Nursing, Level 3, Edmonton Clinic Health Academy, 11405-87 Ave, Edmonton, Alberta, T6G 1C9.

## Abstract

**Introduction:** In Canada, the COVID-19 pandemic has interrupted many routine health services, placed additional strain on the health care system, and resulted in many Canadians being either unable or unwilling to attend routine immunization appointments. We sought to capture and synthesize information about changes to routine immunization programs in response to the pandemic and plans to catch-up any missed immunizations.

**Methods:** Provincial/territorial (P/T) public health leaders were interviewed via teleconference between August-October 2020 to collect information on the following topics: how routine immunization delivery was affected during and after initial lockdown periods, plans to catch-up missed doses, and major challenges and achievements in continuing routine immunization programs. Data were coded and categorized according to common responses and descriptive analysis was performed.

**Results:** Interviews occurred with participants from 11 of 13 P/Ts. School immunization programs were reported to be most negatively affected by the pandemic (*n*=9). In the early pandemic period, infant, preschool, and maternal/prenatal programs were prioritized, with most P/Ts continuing these services with adaptations for COVID-19. After the initial lockdown period, all routine programs were continuing with adaptations in most P/Ts. Infant, preschool, and school programs were most often targeted for catch-up through measures such as appointment rebooking and making additional clinics and/or providers available. Major challenges included resource limitations (e.g., staff shortages, PPE shortages, limited infrastructure) (*n*=11), public health restrictions (*n*=8), and public hesitancy to attend appointments (*n*=5).

**Conclusions:** Canadian routine immunization programs faced some disruptions due to the COVID-19 pandemic, particularly the school, adult, and older adult programs. Further research is needed to determine the measurable impact of the pandemic on routine vaccine coverage levels.

## Introduction

The coronavirus disease 2019 (COVID-19) pandemic has disrupted societies worldwide, resulting in temporary business closures, the cancelling of events, and, central to this study, the interruption of routine immunization programs. In Canada, each province or territory (P/T) is responsible for administering health services, including routine immunizations. The processes for routine vaccine delivery in the P/Ts vary, although most primarily use public health and/or physician delivery (see Appendix 1 for further detail). Due to the pandemic, some public health clinics and physicians’ offices have restricted availability of in-person services, instead offering telephone or online appointments. Furthermore, the pandemic has placed additional strain on the Canadian health care system via COVID-19 testing and contact tracing, as well as health care utilization by COVID-19 patients.^1^ The implementation of public health restrictions has also meant that many Canadians have either been unable or unwilling to attend immunization appointments.^2,3^ As such, despite the recommendation made by the National Advisory Committee on Immunization (NACI) to continue routine immunizations to prevent vaccine-preventable disease outbreaks,^3^ it is likely that Canadian routine immunization programs have been adversely affected by disruptions from the COVID-19 pandemic.

Although Canadian data are not yet available, preliminary data from other jurisdictions have quantified the impact of these disruptions on routine immunization coverage levels since the start of the pandemic. In a global survey, 85% of respondents from 61 countries reported a decline in routine immunization rates in May compared to January and February 2020.^4^ McDonald et al.^5^ found that measles-mumps-rubella (MMR) immunization rates fell 19.8% in England in the three weeks following the introduction of physical distancing measures in late March 2020. Notably, data from New York City showed a 62% decrease in immunization for infants aged <2 years during the early stages of the pandemic (March-April) compared to the same time period in 2019.^6^ It is important to understand and document the processes that led to these drops in immunization coverage, and the strategies that have been undertaken to prevent or recover from them. Identifying the range of strategies, highlighting new and innovative approaches, and learning from successful and unsuccessful approaches will enhance our capacity to respond to similar challenges that we will undoubtedly face in the future.

Little is known about the impact of the COVID-19 pandemic on routine immunization programs in Canadian P/Ts and what measures have been taken by P/T public health officials to meet the challenges posed by the pandemic. As such, the current study was undertaken to capture and synthesize information about the continuity of routine immunization programs during the pandemic through interviews with public health representatives across Canada. We were particularly interested in ascertaining changes to routine immunization programs in response to the pandemic and plans to catch-up any missed immunizations.

## Methods

This pan-Canadian environmental scan involved key informant structured interviews with public health leaders from P/Ts across Canada. The goal was to recruit public health leaders who were knowledgeable of immunization services in all 13 P/Ts. Canadian Immunization Committee members in each P/T were initially targeted for recruitment, but as some individuals were unavailable convenience sampling through referrals from members of the research team, which consists of individuals that partake in immunization research in Canada, P/T Ministries of Health, and the NACI Secretariat at the Public Health Agency of Canada was also used. Key informants were contacted via an initial email sent by the NACI Secretariat, inviting them to participate in the study. Interested individuals were emailed an information sheet and consent form. Informed consent was obtained after the nature of the study and any potential benefits and risks had been fully explained. Up to two email reminders were sent to non-responders to optimize response rate. Some participants were recruited via snowball sampling, with study participants suggesting additional key informants. Interviews (35-60 minutes long) were conducted by members of the research team (HS, AA, MK) from August through October 2020 via teleconference.

The focus of the interviews was to gain in-depth perspectives from individuals who had knowledge about or direct experience with delivery of routine immunization programs during the pandemic. Topics to be explored were identified from scientific literature and news articles, and augmented by the immunization experts on the research team, including the NACI Secretariat. This information was synthesized into questions about: how routine immunization delivery was affected during and after initial lockdown periods, what plans there were to catch-up missed doses, and major challenges and achievements in continuing routine immunization programs. The interview guide was reviewed and edited by immunization experts and pilot tested with an individual who worked in provincial immunization programming, but was not involved in the study, to check face and content validity, flow, and comprehension. Ethical approval for this study was obtained from the Health Research Ethics Board at the University of Alberta.

Interviews were audio-recorded and transcribed verbatim by one member of the research team (HS). The same team member then coded and categorized participant responses. Coding and categorization were validated by other team members (SM, AA) to ensure they accurately reflected and were fully representative of participants’ responses. Descriptive analysis of response counts was performed using Microsoft Excel.

## Results

Invitation emails from the NACI Secretariat were sent to 35 potential participants: 13 agreed to participate, 1 declined, and 21 did not respond. Five more participants were recruited via referrals from other participants. Before the interview, participants were given the opportunity to invite other colleagues to provide additional perspectives on routine immunization programs. Therefore, some interviews contained more than one participant. In total, there were 18 interviews with 25 participants from 11 of the 13 P/Ts. Twelve participants provided a P/T-level perspective, 9 provided a regional/municipal perspective, and 4 provided both P/T and regional/municipal perspectives. Job titles of participants included positions such as Immunization Program or Policy Manager (*n*=7), Medical Officer of Health (*n*=5), and Public Health or Medical Consultant (*n*=3). Table 1 shows the full demographics of the study sample.

**Table 1.** Demographic information of the study sample (N=25).^a^

| **Characteristic** | **Number of participants, *n*** |
| --- | --- |
| **Province/Territory** |  |
| British Columbia | 1 |
| Alberta | 4 |
| Saskatchewan | 3 |
| Manitoba | 4 |
| Ontario | 3 |
| Quebec | 3 |
| Newfoundland and Labrador | 1 |
| Nova Scotia | 3 |
| New Brunswick | 0 |
| Prince Edward Island | 1 |
| Nunavut | 1 |
| Northwest Territories | 1 |
| Yukon | 0 |
| **Perspective** |  |
| Provincial/Territorial | 12 |
| Regional/Municipal | 9 |
| Both | 4 |
| **Job Title** |  |
| Immunization Program or Policy Manager | 7 |
| Medical Officer of Health | 5 |
| Public Health or Medical Consultant | 3 |
| Director of Immunization or Communicable Disease Control | 2 |
| Policy Analyst | 2 |
| Public Health or Communicable Disease Specialist | 2 |
| Other | 4 |

Note. P/T = province/territory; PPE = personal protective equipment
<sup>a</sup>Some P/T responses fell into more than one category

Interview results were synthesized into the following topics, presented below: changes in routine vaccine delivery due to the pandemic; plans for catch-up; and challenges and achievements related to routine immunization program continuity during the pandemic.

### Changes in routine vaccine delivery due to the pandemic

#### Early pandemic period

During the early stages of the pandemic (i.e., March-April 2020, when lockdown measures were first introduced), all P/Ts reported changes to routine immunization programs, with programs being affected to varying degrees. School immunization programs were reported to be the most negatively affected (*n*=9), followed by preschool (*n*=2) and adult (*n*=2) programs. Infant, preschool, and maternal/prenatal programs were most often prioritized. Most P/Ts reported continuing their infant (*n*=11) and preschool programs (*n*=10) with adaptations, such as shortened appointment times (i.e., only providing immunizations) and implementation of COVID-19 public health restrictions (e.g. personal protective equipment [PPE], COVID-19 screening)). Almost half of P/Ts reported a reduction in available appointments for both of these programs (*n*=5). A few P/Ts reported temporary suspension of infant (*n*=3) and preschool (*n*=4) programs, particularly in regions with high COV D-19 incidence. Maternal/prenatal programs were also prioritized, with ten P/Ts commenting that these programs continued with adaptations for public health restrictions, but six also indicating a reduction in appointments. All P/Ts reported suspension of school-based vaccines, due to school closures in mid-March 2020. A few P/Ts reported suspension of adult (*n*=2) and older adult (*n*=3) programs, but overall these programs continued across P/Ts, often with reduced availability and prioritization for high-risk groups. Figure 1a shows all P/T responses for the early pandemic period.

**Figure 1.**
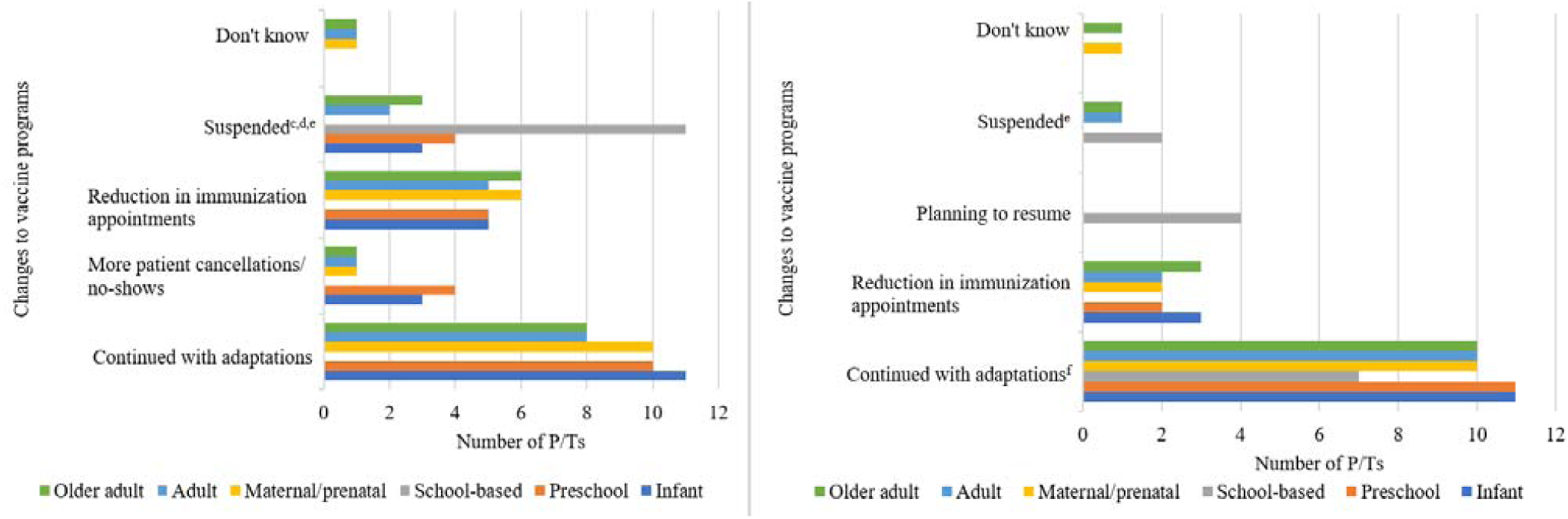
Routine immunization program changes (1a) during initial lockdown period (∼March-April 2020) (left) and (1b) after initial lockdown period (∼May-Oct) (right) (N=11).^a,b^ *Notes*. ^a^See appendix A for list of routine programs in each P/T ^b^Some P/T responses fall into more than one category, so totals may add up to more than 11. ^c^Some P/Ts temporarily suspended the program in some but not all regions. ^d^Some P/Ts continued immunizing high-risk groups only, although programs for the general population were suspended. ^e^In some P/Ts, only non-publicly funded vaccines (e.g., travel vaccines, shingles vaccine) were suspended. ^f^Some P/Ts have resumed vaccination in some but not all regions.

#### Mid-pandemic period

In the period after the initial lockdown (i.e., May-October 2020), most P/Ts reported that infant (*n*=11), preschool (*n*=11), and maternal/prenatal (*n*=10) programs were continuing with adaptations, such as shorter appointment times or implementation of public health restrictions (e.g., PPE, COVID-19 screening). Some P/Ts reported continued suspension of school, adult, and older adult programs, but most P/Ts were continuing these programs with the same adaptations (Figure 1b).

### Plans for routine immunization catch-up

The P/T plans to catch-up vaccines missed due to the pandemic are presented in Tables 2 and 3. School-based vaccines (Table 3) are presented separately, as the strategies for this program are distinct from other programs, which are typically provided in public health clinics and physicians’ offices. For infant and preschool immunizations, many P/Ts reported that clients were contacted to rebook their immunization appointments (Table 2). Some P/Ts increased the number of immunization providers for infant (*n*=4) and preschool (*n*=2) immunizations, some increased the number of clinics, while others were continuing to offer regular infant (*n*=2) and preschool (*n*=3) clinics. Adult and older adult programs were less likely to be actively targeted for catch-up, with P/Ts reporting that these immunizations would be opportunistic at other appointments, provided at regular clinics, or given upon request. Some P/Ts reported increasing providers for adult (*n*=3) and older adult (*n*=2) vaccines.

**Table 2.** Routine infant, pre-school, and adult immunization program catch-up measures as reported by P/Ts (N=11).^a^

| Response | Infant | Preschool | Maternal/<br>prenatal | Adult | Older<br>adult |
| --- | --- | --- | --- | --- | --- |
| **Changes to program delivery** |  |  |  |  |  |
| Increased available providers<br>(e.g., physicians, nurse<br>practitioners, additional nurses) | 4 | 2 | 2 | 3 | 2 |
| Additional clinics offered | 1 | 3 | 0 | 0 | 0 |
| Shortened appointment times | 1 | 1 | 0 | 0 | 0 |
| **Communication** |  |  |  |  |  |
| Clients were contacted and their<br>immunization appointments were<br>rebooked | 5 | 5 | 1 | 2 | 3 |
| Public health released<br>communications to health care<br>providers encouraging them to<br>resume routine immunizations | 1 | 1 | 1 | 1 | 1 |
| **Passive catch-up** |  |  |  |  |  |
| Continuing to offer routine clinics to the best of their ability | 2 | 3 | 0 | 2 | 3 |
| Opportunistic immunization at other appointments | 0 | 0 | 3 | 4 | 5 |
| Clients can be immunized upon request | 0 | 0 | 1 | 1 | 1 |
| No catch-up measures implemented<sup>b</sup> | 2 | 1 | 3 | 2 | 1 |
| Don't know | 0 | 0 | 2 | 2 | 1 |

<sup>a</sup>Some P/T responses fall into more than one category so column totals may add up to more than 11.
<sup>b</sup>Some P/Ts stated that catch-up was not needed for programs that were maintained during the pandemic.

**Table 3.** Routine school-based immunization program catch-up measures as reported by P/Ts (N=11).^a^

| Response | Number of P/Ts (N=11) |
| --- | --- |
| **Summer catch-up program** | **10** |
| Appointment-based | 7 |
| In public health offices | 7 |
| In community (e.g., schools, community centres) | 3 |
| Encouraged providers to offer these vaccines | 2 |
| Don't know | 1 |
| **Fall catch-up program** | **11** |
| Appointment-based | 2 |
| In public health offices | 1 |
| In community (e.g., community centres) | 1 |
| In schools | 10 |
| Use alternative providers (e.g., pharmacists, primary care) | 1 |
| Details of delivery unknown | 1 |

<sup>a</sup>Some P/T responses fall into more than one category so column totals may add up to more than 11.

School-based immunization programs were most likely to be targeted for catch-up, with 10 of 11 P/Ts reporting summer catch-up programs to reach students who had missed immunizations due to school closures, and all P/Ts sharing plans for fall catch-up to reach remaining students (Table 3). The summer catch-up programs were mostly appointment-based (*n*=7) and held in either public health offices (*n*=7) or in community locations, such as schools or community centres (*n*=3). Notably, larger jurisdictions within some P/Ts with high COVID-19 incidence were unable to implement summer catch-up, and instead were planning for fall catch-up. Fall catch-up programs were anticipated by most P/Ts to be held within schools (*n*=10) if restrictions in each jurisdiction allowed it, with appointments at public health offices (*n*=1) or community locations (*n*=1) available for students unable to attend at school.

### Challenges and achievements related to routine immunization program continuity during the pandemic

Many challenges were mentioned by P/Ts in maintaining continuity of routine immunization programs. Most P/Ts indicated that limited resources due to managing the COVID-19 response, including staff shortages, PPE shortages and limited infrastructure, were a significant challenge (*n*=11). Public health restrictions, such as increased infection control measures, school and clinic closures (*n*=8), and public hesitancy to attend clinics/appointments due to COVID-19 (*n*=5) were also frequently reported. Other common challenges included decreased immunization supply due to decreased distribution or changes in delivery schedules (*n*=2), delivery of care including switching to virtual care (*n*=2), limited knowledge and uncertainty about the COVID-19 pandemic (*n*=2), and routine immunizations not being seen as a priority by the public during COVID-19 (*n*=2). Additionally, one P/T mentioned that contacting parents was a challenge.

Despite the many challenges, various achievements were reported by P/Ts in providing routine immunization programs during the pandemic. The most commonly reported were the reorganization of delivery programs to continue immunizations including summer catch-up programs, continuing the immunization of high-risk groups (e.g., immunocompromised, post-exposure prophylaxis), and continuing routine offerings with adaptations (*n*=9). Other frequently reported achievements included greater collaboration between various stakeholders including the public, health services, community agencies, and government (*n*=3), and greater awareness of health care including public appreciation and the importance of immunizations (*n*=3). Achievements indicated by only one P/T were that child immunization rates remained stable and information and technology improvements to immunization registries.

## Discussion

Public health and immunization experts have emphasized the critical need for the continuation of routine immunizations during the COVID-19 pandemic.^2,7,8^ The maintenance of high immunization coverage rates in the population is essential to prevent vaccine-preventable disease outbreaks in the post-pandemic period. For example, multiple measles outbreaks occurred in Guinea in 2015 due to immunization disruptions during the Ebola epidemic.^9^ With COVID-19 lockdown periods causing disruptions to routine immunization, some experts predict that the incidence of diseases such as measles, pertussis, and polio may increase.^10^ It is estimated that at least 80 million children under one year of age are at risk of these diseases due to COVID-19-related disruptions in routine immunization.^11^ Furthermore, missed immunization doses can be problematic as there is evidence that individuals who miss doses may be less likely to catch-up later.^2^ Delaying catch-up of missed doses to the post-pandemic period may place additional strain on a health care system that is already overburdened from the COVID-19 pandemic.^2,12^

Most public health representatives in our study were adamant about the importance of continuing immunization programs, while also following national and provincial guidelines to prevent COVID-19 transmission. Although there were some suspensions of routine immunization programs during the initial lockdown period, most P/Ts eventually resumed all programs with adaptations for COVID-19 restrictions. The suspension of routine immunization programs appears to be in line with global trends.^13^ Of 129 countries for which data were available, 53% reported moderate to severe disruptions or full suspension of immunization between March and April 2020.^11^ In the current study, most P/Ts had catch-up via increasing clinics or providers and contacting clients to rebook appointments for infant, preschool, and school programs. While active catch-up efforts were not implemented for adult and older adult programs in most P/Ts, efforts to immunize these groups at other medical appointments were mentioned. P/Ts appeared to be aligning with Canada’s NACI recommendations for continuity of routine immunizations, with most or all prioritizing immunization for those under two years of age and pregnant women, taking precautions to prevent COVID-19 transmission, and combining vaccinations with other medical visits.^2^

The success in continuing most routine immunization offerings during the pandemic will not be achieved without considerable effort, with P/Ts reporting various challenges, most notably having limited resources due to pandemic response efforts, such as staff shortages, PPE shortages, and limited infrastructure to comply with public health restrictions. Resource shortages have been identified as a major challenge in other countries, with Chandir et al.^14^ reporting a shortage in PPE during the early weeks of the pandemic in Pakistan, and a reduction in the number of available vaccinators due to travel restrictions. Similarly, Ghatak et al.^10^ highlight lack of infrastructure to maintain social distancing and PPE shortages as contributing factors to the reduction in immunization services in India. Given that resource shortages were a challenge for all P/Ts, it may be important to consider factors such as accessing larger spaces for immunization to allow physical distancing and managing immunization programs with fewer staff in any future pandemic preparedness planning. P/Ts also mentioned that public health restrictions made it difficult to deliver routine immunization programs as normal, and that even if programs continued, public hesitancy to attend clinics meant that there was a reduction in attendance. This reduction in attendance aligns with other jurisdictions, with a survey of 400 pediatricians in Italy finding that 31.7% reported a reduction in parents’ compliance with mandatory vaccinations.^12^

Surveys of parents in other jurisdictions have found several barriers to vaccination during the pandemic, including uncertainty around whether vaccination services were operating,^15^ difficulties in making appointments,^15^ and fears of exposure to COVID-19.^15,16^ In particular, public hesitancy to attend immunization clinics has been widely noted as a barrier to providing routine immunizations during the pandemic.^10,12,13,16^ P/Ts in our study also reported that some clients were hesitant to attend immunization appointments, especially during early stages of the pandemic. These findings highlight the importance of clear, consistent, and timely communication from public health or immunization providers regarding the importance of routine immunization and the implementation of measures to prevent disease transmission in clinic settings. A recent study found that advice for parents to continue immunizations resulted in confusion and immunization delays due to conflict with stay-at-home orders.^16^ As well, offering additional hours or days specifically for immunization could encourage individuals to make in-person visits. P/Ts in our study mentioned offering immunizations at other medical appointments, which may be useful for catching up missed adult and older adult immunizations. As noted by MacDonald et al.,^17^ a multi-pronged approach will likely be needed to catch-up missed immunization doses, given the variation in COVID-19 incidence and public health restrictions across P/Ts.

### Strengths and Limitations

A strength of this study was the variety of perspectives that were obtained on routine immunization programs during the COVID-19 pandemic from most P/Ts. As well, the use of key informant interviews rather than survey methods allowed us to gather in-depth perspectives on routine immunization programs in each P/T. However, as only a few select individuals were interviewed from each P/T, the perspectives gathered are not representative of entire P/Ts. Furthermore, there may be variation in individual perspectives across a single P/T, although the perspectives shared were very consistent within a given P/T. At the time the interviews were conducted, most P/Ts did not have up-to-date information on routine immunization coverage rates during the pandemic. Similarly, given that the COVID-19 pandemic is ongoing and there is a lack of available information on immunization coverage rates, we do not know which P/T strategies were the most effective in ensuring continuity of routine immunization programs.

### Implications

This study adds to existing literature by identifying and synthesizing pan-national public health approaches to the continuity of routine immunization programs during the pandemic. Results can inform policymakers and program planners and can assist in future development of national guidelines. As well, we anticipate that the information in this study will enable P/Ts to learn from one another by comparing their approach to maintaining routine immunization during the pandemic with others across Canada.

It will be important to follow up this study with an assessment of whether the planned strategies for routine immunization programs occurred as anticipated. In addition, it will be critical to assess the impact of the COVID-19 pandemic on immunization coverage levels in each P/T. This will permit evaluation of what strategies had the most positive impact on maintenance of immunization coverage levels.

## Conclusion

Our findings show that routine immunization programs across Canadian P/Ts faced some disruptions due to the COVID-19 pandemic, particularly the school, adult, and older adult programs. Catch-up has been implemented for most programs, but there are continued challenges with resource shortages and public health restrictions. Further research is needed to determine the quantitative impact of the pandemic on routine vaccine coverage levels in order to identify potential risks of future vaccine-preventable disease outbreaks.

## Data Availability

Data are not available, due to the potential for participants to be identifiable through the data

## Authors’ Statement

All authors attest that they meet the ICMJE criteria for authorship.

HS – Conceptualization, investigation, data curation, formal analysis, writing (original draft, and review and editing)

AA – Conceptualization, investigation, data curation, supervision, project administration, formal analysis, writing (review and editing)

MD - Conceptualization, methodology, writing (review and editing) ED - Conceptualization, methodology, writing (review and editing)

AG – Conceptualization, methodology, funding acquisition, writing (review and editing) SMeyer – Conceptualization, methodology, writing (review and editing)

JR - Conceptualization, methodology, writing (review and editing) MS – Conceptualization, methodology, writing (review and editing) MT - Conceptualization, methodology, writing (review and editing)

SMacDonald – Conceptualization, methodology, funding acquisition, supervision, formal analysis, writing (original draft, and review and editing)

## Conflict of Interest

MS is supported via salary awards from the BC Children’s Hospital Foundation, the Canadian Child Health Clinician Scientist Program and the Michael Smith Foundation for Health Research. MS has been an investigator on projects funded by GlaxoSmithKline, Merck, Pfizer, Sanofi-Pasteur, Seqirus, Symvivo and VBI Vaccines. All funds have been paid to his institute, and he has not received any personal payments.

## Funding

This work was supported by an operating grant from the Canadian Institutes of Health Research. The funding source had no role in the research or submission process.

## Acknowledgements

This was part of a larger project conducted by the COVImm study team, which included the named authors, as well as: K Benzies; J Bettinger; R Humble; M Kiely; N MacDonald; E Rafferty; and S Wilson. We express particular thanks to M Kiely for her assistance with the interviews and J Libon for her assistance with the literature review and data analysis.

## Appendix 1

**Table A1.** Routine immunization providers by vaccine program and province/territory.

| **Province/**<br>**Territory** | **Infant** (e.g.,<br>diphtheria,<br>rotavirus) | **Preschool** (e.g.,<br>MMRV) | **School-based**<br>(e.g., HPV,<br>meningitis) | **Maternal/Prena**<br>**tal** (e.g.,<br>pertussis, if<br>publicly funded) | **Adult** (e.g., Td<br>booster) | **Older Adult**<br>(e.g.,<br>pneumococcal,<br>shingles) | **Influenza** |
| --- | --- | --- | --- | --- | --- | --- | --- |
| **British**<br>**Columbia** | Public Health (at<br>health units),<br>some family<br>doctors | Public Health (at<br>health units),<br>some family<br>doctors,<br>pharmacists ($\geq 5$<br>years) | Public Health (at<br>schools) | Health units,<br>most pharmacies,<br>some doctor's<br>offices | Health units,<br>most pharmacies,<br>some doctors'<br>offices | Health units,<br>most pharmacies,<br>some doctors'<br>offices | Health units,<br>most pharmacies,<br>some doctor's<br>offices |
| **Alberta** | Public Health<br>(Community or<br>public health<br>centres) | Public Health<br>(Community or<br>public health<br>centres) | Public Health (at<br>schools) | Public health<br>office or<br>pharmacy | Community or<br>public health<br>centres | Community or<br>public health<br>centres, some<br>family doctors,<br>some<br>pharmacists | Community or<br>public health<br>centres,<br>pharmacies ($\geq 5$<br>years), doctor's<br>offices |
| **Saskatchewan** | Public Health<br>clinics | Public Health<br>clinics | Public Health (at<br>schools) | Public health<br>clinics, some<br>doctor and nurse<br>practitioner<br>offices, many<br>pharmacies | Public health<br>clinics, some<br>doctor and nurse<br>practitioner<br>offices,<br>pharmacies | Public health<br>clinics, some<br>doctor and nurse<br>practitioner<br>offices,<br>pharmacies | Public health<br>clinics, some<br>doctor and nurse<br>practitioner<br>offices, many<br>pharmacies ($\geq 5$<br>years) |
| **Manitoba** | Public health<br>offices, nursing<br>stations, doctor's<br>offices,<br>QuickCare | Public health<br>offices, nursing<br>stations, doctor's<br>offices,<br>QuickCare | Public health (at<br>schools) | Public health<br>offices, nursing<br>stations, doctor's<br>offices,<br>pharmacies, | Public health<br>offices, nursing<br>stations, doctor's<br>offices,<br>pharmacies, | Public health<br>offices, nursing<br>stations, doctor's<br>offices,<br>pharmacies, | Local public<br>health office,<br>nursing station,<br>doctor's office,<br>pharmacy ($\geq 7$ |
|  | clinics, and ACCESS Centres | clinics, and ACCESS Centres |  | QuickCare clinics, and ACCESS Centres | QuickCare clinics, and ACCESS Centres | QuickCare clinics, and ACCESS Centres | years), ACCESS Centre, or the nearest immunization clinic |
| **Ontario** | Primary care providers-PHUs in select instances (see note1) | Primary care providers-PHUs in select instances (see note1) | Public health (at schools) | See notes below re universal influenza immunization program (UIIP) providers | Primary care providers-PHUs in select instances | Public health units, community health centres, health care provider offices, pharmacies | Public health units, primary care providers (e.g., physician officers, community health centres, nurse practitioners), occupational health services, pharmacies (≥5 years) |
| **Quebec** | Local community service centres (CLSCs), some doctor's offices | Local community service centres (CLSCs), some doctor's offices | Public health (at schools) | Local community service centres (CLSCs), some doctor's offices, pharmacies | Local community service centres (CLSCs), some doctor's offices, pharmacies | Local community service centres (CLSCs), some doctor's offices, pharmacies | Local community service centres (CLSCs), some doctor's offices, pharmacies (age 6+) |
| **Nova Scotia** | Public health offices, some local primary care providers | Public health offices, some local primary care providers | Public health (at schools) | Public health offices, some local primary care providers, pharmacies | Public health offices, some local primary care providers, pharmacies | Public health offices, some local primary care providers, pharmacies | Public health offices, some local primary care providers, pharmacies |
| **Newfoundland** | Local health units | Local health units | Public health (at schools) | Public health nursing clinics only | Public health | Public health | Public health, healthcare occupational health services and physician's offices |
| **New Brunswick** | Public health clinics | Public health clinics | Public health (at schools) | Family physicians, nurse practitioners, pharmacists, midwives | Public health clinics, family physicians, nurse practitioners | Family physicians, nurse practitioners, pharmacists | Public Health Offices (children 6-59 months), some pharmacies (5+ years), family physicians, midwives (pregnant women) and other health care providers |
| **Prince Edward Island** | Public health nursing offices | Public health nursing offices | Public health (at schools) | Public Health Nursing office | Local Health PEI Public Health Nursing office, family doctor or nurse practitioner's office or a pharmacy, depending on the vaccine | Local Health PEI Public Health Nursing office, family doctor or nurse practitioner's office or a pharmacy, depending on the vaccine | Family physicians, nurse practitioners, pharmacies, and Health PEI Flu Clinics |
| **Yukon** | Community health centres | Community health centres | Public health (at schools) | Community health centres | Community health centres | Community health centres | Flu clinics at community health centres |
| **Northwest Territories** | Health centre, public health unit | Health centre, public health unit | Public health (at schools) | Health centre, public health unit | Health centre, public health unit | Health centre, public health unit | Health centre, public health unit |
| **Nunavut** | Community health centres and public health offices | Community health centres and public health offices | Public health (at schools) | Community health centres and public health offices | Community health centres and public health offices | Community health centres and public health offices | Community health centres and public health offices |

## Notes

### Competing Interest Statement

MS is supported via salary awards from the BC Childrens Hospital Foundation, the Canadian Child Health Clinician Scientist Program and the Michael Smith Foundation for Health Research. MS has been an investigator on projects funded by GlaxoSmithKline, Merck, Pfizer, Sanofi-Pasteur, Seqirus, Symvivo and VBI Vaccines. All funds have been paid to his institute, and he has not received any personal payments.

### Author Declarations

University of Alberta Health Research Ethics Board

## References

1. Barrett K, Khan YA, Mac S, Ximenes R, Naimark DMJ, Sander B. Estimation of COVID-19 induced depletion of hospital resources in Ontario, Canada. CMAJ. 2020;192:E640–6.

2. Ontario Ministry of Health. Guidance for immunization services during COVID-19 [Internet]. Ottawa: Ontario Ministry of Health; 2020 [updated 2020 Aug 25; cited 2021 Jan 14]. Available from: http://www.health.gov.on.ca/en/pro/programs/publichealth/coronavirus/docs/Immunization_Services_during_COVID-19_08-26-2020.pdf

3. National Advisory Committee on Immunization. Interim guidance on continuity of immunization programs during the COVID-19 pandemic [Internet]. Government of Canada; 2020 [updated 2020 May 13; cited 2020 Jan 14]. Available from: https://www.canada.ca/en/public-health/services/immunization/national-advisory-committee-on-immunization-naci/interim-guidance-immunization-programs-during-covid-19-pandemic.html

4. World Health Organization. Special feature: immunization and COVID-19 [Internet]. World Health Organization; 2020 [cited 2021 Jan 18]. Available from: https://www.who.int/immunization/monitoring_surveillance/immunization-and-covid-19/en/

5. McDonald HI, Tessier E, White JM, Woodruff M, Knowles C, Bates C, et al. Early impact of the coronavirus disease (COVID-19) pandemic and physical distancing measures on routine childhood vaccinations in England, January to April 2020. Euro Surveill. 2020;25:1–6.

6. Langdon-Embry M, Papadouka V, Cheng I, Almashhadani M, Ternier A, Zucker JR. Rebound in routine childhood vaccine administration following decline during the COVID-19 pandemic —New York City, March 1–June 27, 2020. Morb Mortal W kly Rep. 2020;69:999–1001.

7. Bonanni P, Angelillo IF, Villani A, Biasci P, Scotti S, Russo R, et al. Maintain and increase vaccination coverage in children, adolescents, adults and elderly people: Let’s avoid adding epidemics to the pandemic: Appeal from the Board of the Vaccination Calendar for Life in Italy: Maintain and increase coverage also by re-organizing vaccination services and reassuring the population. Vaccine. 2021;39(8):1187–9.

8. World Health Organization. Guidance on routine immunization services during COVID-19 pandemic in the WHO European Region [Internet]. Copenhagen (DK): World Health Organization; 2020 [cited 2021 Feb 19]. 6 p. Available from: https://apps.who.int/iris/bitstream/handle/10665/334123/WHO-EURO-2020-1059-40805-55114-eng.pdf

9. Suk JE, Jimenez AP, Kourouma M, Derrough T, Baldé M, Honomou P, et al. Post-Ebola measles outbreak in Lola, Guinea, January–June 2015. Emerg Infect Dis. 2016;22(6):1106–8.

10. Ghatak N, Marzo RR, Saleem SM, Sharma N, Bhattacharya S, Singh A. Impact on routine immunization services during the lockdown period in India: Implications and future recommendations. Eur J M ol Clin Med. 2020;7(5):35–40.

11. World Health Organization. At least 80 million children under one at risk of diseases such as diphtheria, measles and polio as COVID-19 disrupts routine vaccination efforts, warn Gavi, WHO and UNICEF [Internet]. Geneva: World Health Organization; 2020 May 22 [cited 2021 Mar 2]. Available from: https://www.who.int/news/item/22-05-2020-at-least-80-million-children-under-one-at-risk-of-diseases-such-as-diphtheria-measles-and-polio-as-covid-19-disrupts-routine-vaccination-efforts-warn-gavi-who-and-unicef

12. Bechini A, Garamella G, Giammarco B, Zanella B, Flori V, Bonanni P, et al. Paediatric activities and adherence to vaccinations during the COVID-19 epidemic period in Tuscany, Italy: a survey of paediatricians. J Prev M ed Hyg. 2020;61:E125–9.

13. Dinleyici EC, Borrow R, Safadi MAP, van Damme P, Munoz FM. Vaccines and routine immunization strategies during the COVID-19 pandemic. Hum Vaccin Immunother. 2021;17(2):400–7.

14. Chandir S, Siddiqi DA, Mehmood M, Setayesh H, Siddique M, Mirza A, et al. Impact of COVID-19 pandemic response on uptake of routine immunizations in Sindh, Pakistan: An analysis of provincial electronic immunization registry data. Vaccine. 2020;38:7146–55.

15. Bell S, Clarke R, Paterson P, Mounier-Jack S. Parents’ and guardians’ views and experiences of accessing routine childhood vaccinations during the coronavirus (COVID-19) pandemic: A mixed methods study in England. PLoS One. 2020;15(12):e0244049.

16. Alsuhaibani M, Alaqeel A. Impact of the COVID-19 pandemic on routine immunization in Saudi Arabia. Vaccines. 2020;8:581.

17. MacDonald NE, Comeau JL, Dubé È, Bucci LM. COVID-19 and missed routine immunizations: designing for effective catch-up in Canada. Can J Public Health. 2020;111:469–72.

